## Supplemental files 1, 2, 3 and 4 for "The Sexual and Reproductive Health needs and preferences of youths in sub-Saharan Africa: A meta-synthesis": S1_file.pdf

**List of countries in sub-Saharan Africa**

Angola, Benin, Botswana, Burkina Faso, Burundi, Cameroon, Cape Verde, Central African Republic, Chad, Comoros, Republic of the Congo, Democratic Republic of the Congo, Cote d'Ivoire, Equatorial Guinea, Eritrea, Ethiopia, Gabon, Gambia, Ghana, Guinea, Guinea-Bissau, Kenya, Liberia, Madagascar, Malawi, Mali, Mauritania, Mauritius, Mozambique, Namibia, Niger, Nigeria, Rwanda, Sao Tome and Principe, Senegal, Seychelles, Sierra Leone, South Africa, South Sudan, Swaziland, Tanzania, Togo, Uganda, Zambia, Zimbabwe and Lesotho.
