## Supplemental files 1, 2, 3 and 4 for "The Sexual and Reproductive Health needs and preferences of youths in sub-Saharan Africa: A meta-synthesis": S2_file.pdf

### Search histories

**Ovid Medline**

Ovid MEDLINE(R) and Epub Ahead of Print, In-Process, In-Data-Review & Other Non-Indexed Citations, Daily and Versions(R) <1946 to June Week 4 2023>

1 Young pe*.mp.

2 Young Adult/

3 youth.mp.

4 adolescen*.mp.

5 teen*.mp.

6 Child/

7 1 or 2 or 3 or 4 or 5 or 6

8 Sexual Health/

9 Reproductive health*.mp.

10 8 or 9

11 service*.mp.

12 Clinic*.mp.

13 Program*.mp.

14 Education/

15 Counsel*.mp.

16 Promot*.mp. 1247350

17 11 or 12 or 13 or 14 or 15 or 16

18 10 and 17

19 sub-Saharan Africa.mp.

20 Angola/

21 Benin/

22 Botswana/

23 Burkina Faso/

24 Burundi/

25 Cameroon/

26 Cape Verde.mp.

27 Central African Republic/

28 Chad/

29 Comoros/

30 Republic of the Congo.mp.

31 "Democratic Republic of the Congo"/

32 Cote d'Ivoire/

33 Equatorial Guinea/

34 Eritrea/

35 Ethiopia/

36 Gabon/

37 Gambia/

38 Ghana/

39 Guinea/

40 Guinea-Bissau/

41 Kenya/

42 Liberia/

43 Madagascar/

44 Malawi/

45 Mali/

46 Mauritania/

47 Mauritius/

48 Mozambique/

49 Namibia/

50 Niger/

51 Nigeria/

52 Rwanda/

53 Senegal

54 Seychelles/

55 Sierra Leone/

56 South Africa/

57 South Sudan/

58 Swaziland.mp.

59 Tanzania/

60 Togo/

61 Uganda/

62 Zambia/

63 Zimbabwe/

64 Lesotho/

65 Sao Tome*.mp.

66 19 or 20 or 21 or 22 or 23 or 24 or 25 or 26 or 27 or 28 or 29 or 30 or 31 or 32 or 33 or 34 or 35 or 36 or 37 or 38 or 39 or 40 or 41 or 42 or 43 or 44 or 45 or 46 or 47 or 48 or 49 or 50 or 51 or 52 or 53 or 54 or 55 or 56 or 57 or 58 or 59 or 60 or 61 or 62 or 63 or 64 or 6

67 7 and 18 and 66

**CINAHL plus**

S67 S7 AND S18 AND S66

S66 S19 OR S20 OR S21 OR S22 OR S23 OR S24 OR S25 OR S26 OR S27 OR S28 OR S29 OR S30 OR S31 OR S32 OR S33 OR S34 OR S35 OR S36 OR S37 OR S38 OR S39 OR S40 OR S41 OR S42 OR S43 OR S44 OR S45 OR S46 OR S47 OR S48 OR S49 OR S50 OR S51 OR S52 OR S53 OR S54 OR S55 OR S56 OR S57 OR S58 OR S59 OR S60 OR S61 OR S62 OR S63 OR S64 OR S65

S65 (MH "Lesotho")

S64 (MH "Zimbabwe")

S63 (MH "Zambia")

S62 (MH "Uganda")

S61 (MH "Togo")

S60 (MH "Tanzania")

S59 (MH "Swaziland")

S58 "south sudan"

S57 (MH "South Africa")

S56 (MH "Sierra Leone")

S55 "Seychelles"

S54 (MH "Senegal")

S53 "Sao Tome and Principe"

S52 (MH "Rwanda")

S51 (MH "Nigeria")

S50 (MH "Niger")

S49 (MH "Namibia")

S48 (MH "Mozambique")

S47 "Mauritius"

S46 (MH "Mauritania")

S45 (MH "Mali")

S44 (MH "Malawi")

S43 (MH "Madagascar")

S42 (MH "Liberia")

S41 (MH "Kenya")

S40 (MH "Guinea-Bissau")

S39 (MH "Guinea")

S38 (MH "Ghana")

S37 (MH "Gambia")

S36 (MH "Gabon")

S35 (MH "Ethiopia")

S34 (MH "Eritrea")

S33 (MH "Equatorial Guinea")

S32 (MH "Cote d'Ivoire")

S31 (MH "Democratic Republic of the Congo")

S30 "Republic of the Congo"

S29 "Comoros"

| | |
|---|---|
| S28 | (MH "Chad") |
| S27 | (MH "Central African Republic") |
| S26 | (MH "Cape Verde") |
| S25 | (MH "Cameroon") |
| S24 | (MH "Burundi") |
| S23 | (MH "Burkina Faso") |
| S22 | (MH "Botswana") |
| S21 | (MH "Benin") |
| S20 | (MH "Angola") |
| S19 | "sub-saharan Africa" |
| S18 | S10 AND S17 |
| S17 | S11 OR S12 OR S13 OR S14 OR S15 OR S16 |
| S16 | "Promot*" |
| S15 | "Counsel*" |
| S14 | (MH "Education") |
| S13 | "Program*" |
| S12 | "Clinic*" |
| S11 | "Service*" |
| S10 | S8 OR S9 |
| S9 | (MH "Reproductive Health") |
| S8 | (MH "Sexual Health") |
| S7 | S1 OR S2 OR S3 OR S4 OR S5 OR S6 |
| S6 | (MH "Child") |
| S5 | "Teen*" |
| S4 | "Youth" |
| S3 | "Adolescen*" |
| S2 | (MH "Young Adult") |
| S1 | "Young pe*" |

**PsychINFO**

Database: APA PsycInfo <1806 to June Week 4 2023>

Search Strategy:

--------------------------------------------------------------------------------

1 Young pe*.mp.

2 Young adult.mp.

3 Youth.mp.

4 Adolescen*.mp.

5 teen*.mp.

6 child.mp.

7 1 or 2 or 3 or 4 or 5 or 6

8 exp Sexual Health/

9 Reproductive health*.mp.

10 8 or 9

11 service*.mp.

12 clinic*.mp.

13 Program*.mp.

14 exp Education/

15 Counsel*.mp.

16 promot*.mp.

17 11 or 12 or 13 or 14 or 15 or 16

18 10 and 17

19 sub-Saharan Africa.mp.

20 Angola.mp.

21 Benin.mp.

22 Botswana.mp.

23 Burkina Faso.mp.

24 Burundi.mp.

25 Cameroon.mp.

26 Cape Verde.mp.

27 Central African Republic.mp.

28 Chad.mp.

29 Comoros.mp.

30 Republic of the Congo.mp.

31 Democratic Republic of the Congo.mp.

32 Cote d'Ivoire.mp.

33 Equatorial Guinea.mp.

34 Eritrea.mp.

35 Ethiopia.mp.

36 Gabon.mp.

37 Gambia.mp.

38 Ghana.mp.

39 Guinea.mp.

40 Guinea-Bissau.mp.

41 Kenya.mp.

42 Liberia.mp.

43 Madagascar.mp.

44 Malawi.mp.

45 Mali.mp.

46 Mauritania.mp.

47 Mauritius.mp.

48 Mozambique.mp.

49 Namibia.mp.

50 Nigeria.mp.

51 Niger.mp.

52 Rwanda.mp.

53 (Sao Tome and Principe).mp. [mp=title, abstract, heading word, table of contents, key concepts, original title,

tests & measures, mesh word]

54 Senegal.mp.

55 Seychelles.mp.

56 Sierra Leone.mp.

57 South Africa.mp.

58 South Sudan.mp.

59 Swaziland.mp.

60 Tanzania.mp.

61 Togo.mp.

62 Uganda.mp.

63 Zambia.mp.

64 Zimbabwe.mp.

65 Lesotho.mp.

66 19 or 20 or 21 or 22 or 23 or 24 or 25 or 26 or 27 or 28 or 29 or 30 or 31 or 32 or 33 or 34 or 35 or 36 or 37 or

38 or 39 or 40 or 41 or 42 or 43 or 44 or 45 or 46 or 47 or 48 or 49 or 50 or 51 or 52 or 53 or 54 or 55 or 56 or 57 or

58 or 59 or 60 or 61 or 62 or 63 or 64 or 65

67 7 and 18 and 66
