## Supplemental files 1, 2, 3 and 4 for "The Sexual and Reproductive Health needs and preferences of youths in sub-Saharan Africa: A meta-synthesis": S3_Table.pdf

### Data extraction summary on youth's needs and preferences regarding SRHS in sub-Saharan Africa

| S/N | Tittle | Authors/ Publication date | Country of study | Name of Journal/impact factor | Context | Design | Target adolescent population | Method of data collection | Emergent themes |
|---|---|---|---|---|---|---|---|---|---|
| 1. | Addressing the sexual and reproductive health needs of young adolescents living with HIV in South Africa | Vujovic, M., Struthers, H., Meyersfeld, S., Dlamini, K. and Mabizela, N (2014) | South Africa | Children and youth services review (ELSEVIER)/ 2.519 | facility based | Qualitative (no mention of the specific qualitative design) | Male and female living with HIV (ALWHIV) 10-14years | Focus group discussion (FGD) | 1. Service provision<br>2. Information, education, and communication<br>3. Risk perception and management |
| 2. | The extent to which the design of available reproductive health interventions fit the reproductive health needs of adolescents living in urban poor settings of Kisenyi, Kampala, Uganda | Tuhebwe, D., Babirye, S., Ssendagire, S. and Ssengooba, F. (2021) | Uganda | BMC Public health/ 4.135 (2-year impact) | Community based | Qualitative (specific qualitative design not mentioned) | Male and female aged 15-19years | FGD | 1.Sexual reproductive health (SRH) needs<br>2. Social needs.<br>3. Extent of fit between intervention design features and priority reproductive health needs of the adolescents |
| 3. | What do South African adolescents want in a sexual health service? Evidence from the South African Studies on HIV in Adolescents (SASHA) project | Smith, P., Marcus, R., Bennie, T., Nkala, B., Nchabeleng, M., Latka, M.H., Gray, G., Wallace, | South Africa | South African Medical Journal (SAMJ)/1.500 | community based | Qualitative (specific qualitative design not mentioned) | Male and female aged 12-17years | FGD | 1. The need for the provision of dedicated adolescent health services.<br>2. Tailored services with developmentally appropriate information.<br>3. Emphasis on the desire for confidentiality and trusting |

| | | | | | | | | | |
|---|---|---|---|---|---|---|---|---|---|
| | | M. and Bekker, L.G. (2018) | | | | | | | relationships with health-care staff.<br>4. Availability of services and information |

| S/N | Tittle | Authors/ publication date | Country of study | Name of Journal/impact factor | Context | Design | Target adolescent popula | Method of data collection | Themes |
|---|---|---|---|---|---|---|---|---|---|
| 4. | Engaging young people in the design of a sexual reproductive health intervention: Lessons learnt from the Yathu Yathu ("For us, by us") formative study in Zambia | Simuyaba, M., Hensen, B., Phiri, M., Mwansa, C., Mwenge, L., Kabumbu, M., Belemu, S., Shanaube, K., Schaap, A., Floyd, S. and Fidler, S. (2021) | Zambia | BMC Health Services Research (ELSEVIER)/2.908 (2-year impact) | Community based | Participatory qualitative research | Male and female, 15-24 years | FGD, observation and In-depth interviews (IDIs) | 1. Community mapping: context of adolescents and young people (AYP)'s SRH<br>2. Primary discussion<br>3. Consolidation of AYP's views |
| 5. | Youth accessing reproductive health services in Malawi: drivers, barriers, and suggestions from the perspectives of youth and parents. | , Self A., Chipokosa, S., Misomali, A., Aung, T., Harvey, S.A., Chimchere, M., Chilembwe, J., Park, L., Chalimba, C., Monjeza, E. and Kachale, F. (2018) | Malawi | Reproductive Health (BMC) /3.355 (2-year impact) | Facility based | Qualitative (specific qualitative design not mentioned) | Male and female aged 15-24years | FGD | 1. Drivers of youth accessing family planning services<br>2. Barriers' youth face accessing family planning services Misconceptions and perceived side-effects<br>3. Suggestions from participants for improving family planning services |
| 6. | Young people's perceptions of youth-oriented health services in urban Soweto, South Africa: a qualitative investigation | Schriver, B., Meagley, K., Norris, S., Geary, R. and Stein, A.D. (2014) | South Africa | Health Services Research (BMC)2.908 (2-year impact) | community based. | Ground theory as described by Borgatti and Strauss and Corbin. | Male and female youths aged 21-22years | IDIs | 1. Perception of current health services.<br>2. Knowledge of youth friendly services.<br>3. Attitudes toward alternative health services. |

| S/N | Tittle | Authors/ publication date | Country of study | Name of Journal/impact factor | Context | Design | Target adolescent population | Method of data collection | Themes |
|---|---|---|---|---|---|---|---|---|---|
| 7. | Provision of Reproductive Health Services for Adolescents -- Report of a Study in Two Local Government Areas (LGAs) of Nigeria | Olukoya, A. (1996) | Nigeria | Early Child Development and Care (Taylor & Francis)/1.206(2-year impact) | Facility based | Mixed study (Specific qualitative design not mentioned) | Out of school and in-school Male and female adolescents. Authors stated ''less than 15years'' | FGD and direct observation | 1. Perceptions of the health problems of adolescents.<br>2. Help-seeking behaviour. |
| 8. | Sexual and reproductive health services (SRHS) for adolescents in Enugu state, Nigeria: a mixed methods approach. | Odo, A.N., Samuel, E.S., Nwagu, E.N., Nnamani, P.O. and Atama, C.S. (2018) | Nigeria | Health Services Research (BMC)2.908 (2-year impact) | Community based | Mixed (Specific qualitative design not mentioned) | Male and female aged 12-22years | Interview (type-unclear) and FGD | 1. Availability of SRHS for Adolescents.<br>2. Accessibility of SRHS to adolescents. |
| 9. | Access to information and use of adolescent sexual reproductive health services: Qualitative exploration of barriers and facilitators in Kisumu and Kakamega, Kenya | Mutea, L., Ontiri, S., Kadiri, F., Michielesen, K. and Gichangi, P.(2020) | kenya | PLOS ONE (No impact factor documentation on the journal's home page) | Community based. | Qualitative (specific qualitative design not mentioned) | Male and female adolescents aged 15-19 years | FGD, IDIs and Key Informant interviews (KIIs) | 1. Common issues perceived to affect the health of adolescents in the community.<br>2. Barriers to access and use of SRH information and services by adolescents.<br>3. Facilitators to access and use of ASRH information |

| | | | | | | | | | |
|---|---|---|---|---|---|---|---|---|---|
| | | | | | | | | | and services. |

| S/ N | Tittle | Authors/ publication date | Country of study | Name of Journal/impact factor | Context | Design | Target adolescent population | Method of data collection | Themes |
|---|---|---|---|---|---|---|---|---|---|
| 10 | Does Making Clinic-based Reproductive Health Services More Youth-friendly Increase Service Use by Adolescents? Evidence From Lusaka, Zambia | Mmari, K.N. and Magnani, R.J. (2003) | Zambia | Journal of Adolescent Health (ELSEVIER)/7.83 | Facility based | Mixed study (Specific qualitative design not mentioned) | Male and female aged 15-24years. | FGDs and interviews | 1. Youth-Friendliness of Health Service. |
| 11. | Adolescents living with HIV in the Copperbelt Province of Zambia: Their reproductive health needs and experiences. | McCarraher, D.R., Packer, C., Mercer, S., Dennis, A., Banda, H., Nyambe, N., Stalter, R.M., Mwansa, J.K., Katayamoyo, P. and Denison, J.A (2018) | Zambia | PLOS ONE | Facility based | Mixed (Specific qualitative design not mentioned) | Male and female aged 15–18years | IDIs | 1. Sexual experiences including forced sex.<br>2. Sex partner age and HIV Disclosure.<br>3. Contraceptive use.<br>4. Fertility desires and prevention of mother-to-child transmission of HIV. |
| 12. | Rights-based services for adolescents living with HIV: adolescent self-efficacy and implications for health systems in Zambia | Mburu, G., Hodgson, I., Teltschik, A., Ram, M., Haamujompa, C., Bajpai, D. and Mutali (2013) | Zambia | Reproductive Health Matters (Taylor & Francis)5.732 | Facility based. | Qualitative (specific qualitative design not mentioned) | Male and female adolescents aged 10-19years | Semi-structured interviews and FGDs | 1. A sense of rights, entitlement, and expectation.<br>2. Expressing unmet need. |

| S/N | Tittle | Authors/ publication date | Country of study | Name of Journal/impact factor | Context | Design | Target adolescent population | Method of data collection | Themes |
|---|---|---|---|---|---|---|---|---|---|
| 13. | Accessing Sexual and Reproductive Health Information and Services: A Mixed Methods Study of Young Women's Needs and Experiences in Soweto, South Africa | Lince-Deroche, N., Hargey, A., Holt, K. and Shochet, T. (2015) | South Africa | African Journal of Reproductive Health. (It appears, there is no impact factor documentation on the journal website) | Facility based | Mixed (Grounded theory used for the qualitative approach) | Female adolescents 18-24 years | Semi-structured interviews. | 1. Contraception: knowledge and access to services<br>2. Abortion services.<br>3. HIV testing and condom Use.<br>4. Gender-based violence.<br>5. Concerns and support systems |
| 14. | Adolescents' Reproductive Health Problems, Service Preferences, and Accessibility. | Kimo, K. and Makuria, K. (2017) | Ethiopia | Pakistan Journal of Psychological Research (No impact factor seen) | Facility based | Mixed (Specific qualitative design not mentioned) | Male and female adolescents (12-19years) | FGD | Reproductive health service accessibility (the only qualitative theme). |
| 15 | Living as an adolescent with HIV in Zambia – lived experiences, sexual health, and reproductive needs. | Hodgson I, Julia Ross, Choolwe Haamujompa & D. Gitau-Mburu (2012) | Zambia | AIDS Care-Psychological and Socio-Medical Aspects of AIDS/HIV (Taylor & Francis) (No impact factor seen) | Facility based | Qualitative (specific qualitative design not mentioned) | Male and female adolescents aged 10-19years | semi-structured interviews and FGD | 1. Informational needs.<br>2. Psychosocial and disclosure needs.<br>3. Sexual and reproductive health (SRH) needs of adolescents.<br>4. Health systems and services capacity. |

| S/N | Tittle | Authors/ publication date | Country of study | Name of Journal/ impact factor | Context | Design | Target adolescent population | Method of data collection | Themes |
|---|---|---|---|---|---|---|---|---|---|
| 16 | Young people's perception of sexual and reproductive health services in Kenya | Godia, P.M., Olenja, J.M., Hofman, J.J. and Van Den Broek, N. (2014) | Kenya | Health Services Research (BMC)2.908 (2-year impact) | Facility based | Qualitative (Specific qualitative design not mentioned) | Male and female aged 10–24 years. | FGDs and semi-structured in-depth interviews (IDIs) | 1. SRH problems faced by young people.<br>2. Addressing the SRH needs of young people.<br>3. Perceptions of existing SRH services.<br>4. Suggestions on how to improve SRH services |
| 17. | Understanding sexual and reproductive health needs of adolescents: evidence from a formative evaluation in Wakiso district, Uganda | Atuyambe, L.M., Kibira, S.P., Bukenya, J., Muhumuza, C., Apolot, R.R. and Mulogo, E. (2015) | Uganda | Reproductive Health (BMC) /3.355 (2-year impact) | Community based | Qualitative (Specific qualitative design not mentioned) | Male and female aged 10-19years | FGD | 1. Main adolescent health problems.<br>2. Adolescent SRH needs.<br>3. Health seeking behaviour and attitudes towards services. |
| 18. | Adolescent human immunodeficiency virus self-management: Needs of adolescents in the Eastern Cape | Adams, L. and Crowley, T. (2021) | South Africa | African Journal of Primary Health Care & Family Medicine /1.04 | Facility based. | qualitative exploratory-descriptive research design | Male and female adolescents aged 14 -19 years | Semi-structured interview | 1. Knowledge of human immunodeficiency virus and sexual reproductive health.<br>2. Self-regulation skills.<br>3. Self-management resources. |
| 19 | Preferences for accessing sexual and reproductive health | Adhiambo, H. F, Ngayo, M. and | Kenya | PLOS ONE | Facility based | Qualitative (Specific qualitative | Male and female adolescents | IDIs and FGDs | 1. Preferences of venue for receiving SRH services. |

| | | | | | | | | | |
|---|---|---|---|---|---|---|---|---|---|
| | services among adolescents and young adults living with HIV/ AIDs in Western Kenya: A qualitative study | Kwena, Z. (2022) | | | | design not mentioned) | aged 14 -24 years | | 2. Preferences of qualities of SRH counsellors. |
| 20 | Pregnancy and STI/HIV prevention intervention preferences of South African adolescent girls: findings from a cultural consensus modelling qualitative study | Twitty et al. (2023) | South Africa | Culture, Health & Sexuality An International Journal for Research, Intervention and Care (Taylor & Francis) | School-based | Qualitative (Specific qualitative design not mentioned) | Females aged between 14 and 17 years | Semi-structured interviews | 1. Intervention content<br>2. Intervention delivery format<br>3. Intervention setting |
