## Supplemental files 1, 2, 3 and 4 for "The Sexual and Reproductive Health needs and preferences of youths in sub-Saharan Africa: A meta-synthesis": S4_Table.pdf

**S4 - CASP appraisal of included papers on the needs and preferences of youths regarding sexual and reproductive health services in sub-Saharan Africa**

| S/N | Title | Author (s)/Date of publication | Responses and scoring of the 10 questions on the CASP checklist | | | | | | | | | | Judgement/ Total | Reason |
|---|---|---|---|---|---|---|---|---|---|---|---|---|---|---|
| | | | Q1 | Q2 | Q3 | Q4 | Q5 | Q6 | Q7 | Q8 | Q9 | Q10 | | |
| 1 | Addressing the SRH needs of young people living with HIV in South Africa | Vujovic et al. (2014) | Yes | Yes | Yes | Can't tell | Yes | Can't tell | Yes | Yes | Yes | Valuable resource | | There was a lack of clarity regarding the authors' position and recruitment process. However, the study provides a valuable resource since it targeted the SRH preferences of HIV-positive adolescents. |
| | | | 2 | 2 | 2 | 1 | 2 | 1 | 2 | 2 | 2 | 2 | 18 | |
| . 2 | The extent to which the design of available reproductive health interventions fit the reproductive health needs of adolescents living in urban poor settings of Kisenyi, Kampala, Uganda. | Tuhebwe et al. (2021) | Yes | Yes | Yes | Yes | Yes | Yes | Yes | Yes | Yes | Valuable resource | | The study revealed a gap between adolescents' RH needs and the RH interventions in Kisenyi slum. Policy makers may be able to use this information to design appropriate interventions that would consider adolescents' livelihood and sanitation needs. |
| | | | 2 | 2 | 2 | 2 | 2 | 2 | 2 | 2 | 2 | 2 | 20 | |
| 3. | What do South African adolescents want in a sexual health service? Evidence from the South African Studies on HIV in adolescents (SASHA) project | Smith et al. (2018) | Yes | Yes | Yes | Yes | Yes | Can't tell | Yes | Yes | Yes | Valuable resource | | Although the authors did not make clear their own role, potential bias and how participant or researcher's distress were addressed, the study is valuable it revealed what services adolescents preferred and this could be helpful in future design of SRHS for young people |
| | | | 2 | 2 | 2 | 2 | 2 | 1 | 2 | 2 | 2 | 2 | 19 | |

| S/N | Title | Author (s)/Date of publication | Responses and scoring of the 10 questions on the CASP checklist | | | | | | | | | | Judgement/ Total | Reason |
|---|---|---|---|---|---|---|---|---|---|---|---|---|---|---|
| | | | Q1 | Q2 | Q3 | Q4 | Q5 | Q6 | Q7 | Q8 | Q9 | Q10 | | |
| 4 | Engaging young people in the design of a sexual reproductive health intervention: Lessons learnt from the Yathu Yathu ("For us, by us") formative study in Zambia | Simuyaba et al. (2021) | Yes | Yes | Yes | Yes | Yes | Yes | Yes | Yes | Yes | Valuable resource | | Through participatory research, this study identified gaps in SRH provision to adolescents and engaged adolescents to design the kind of SRH services they wanted. |
| | | | 2 | 2 | 2 | 2 | 2 | 2 | 2 | 2 | 2 | 2 | 20 | |
| 5. | Youth accessing reproductive health services in Malawi: drivers, barriers, and suggestions from the perspectives of youth and parents. | Self et al. (2018) | Yes | Yes | Yes | Yes | Yes | Can't tell | Yes | Yes | Yes | Valuable resource | | Although, it is unclear if there was any event during the study and how it was handled including the author's position, the study highlighted the need for re-orientation of adolescent friendly SRH services to include programming and policy development. |
| | | | 2 | 2 | 2 | 2 | 2 | 1 | 2 | 2 | 2 | 2 | 19 | |
| 6. | Young people's perceptions of youth-oriented health services in urban Soweto, South Africa: a qualitative investigation | Schriver et al. (2014) | Yes | Yes | Yes | Yes | Yes | yes | Yes | Yes | Yes | Valuable resource | | The study illuminated the gaps between youth knowledge and what young people in Soweto receive in terms of SRHS. |
| | | | 2 | 2 | 2 | 2 | 2 | 2 | 2 | 2 | 2 | 2 | 20 | |

| S/N | Title | Author (s)/Date of publication | Q1 | Q2 | Q3 | Q4 | Q5 | Q6 | Q7 | Q8 | Q9 | Q10 | Judgement/ Total | Reason |
|---|---|---|---|---|---|---|---|---|---|---|---|---|---|---|
| | | | **Responses and scoring of the 10 questions on the CASP checklist** | | | | | | | | | | | |
| 7 | Provision of Reproductive Health Services for Adolescents -- Report of a Study in Two Local Government Areas (LGAs) of Nigeria | Olukoya, A. (2021) | Yes | Yes | Yes | Can't tell | Can't tell | No | No | Can't tell | Can't tell | Less valuable | | Although it appeared that the study was deficient in documentation including ethical procedures, participant recruitment and data collection process, it is somewhat valuable because gaps in SRH service provision to adolescents were highlighted. This could assist in the design of interventions to meet the needs of adolescents. |
| | | | 2 | 2 | 2 | 1 | 1 | 0 | 0 | 1 | 1 | 1 | **11** | |
| 8. | Sexual and reproductive health services (SRHS) for adolescents in Enugu state, Nigeria: a mixed methods approach. | Odo et al. (2018) | Yes | Yes | Yes | Yes | Yes | Can't tell | Yes | Can't tell | Yes | Valuable resource | | The study showed that most SRH services were accessible geographically. There were, however, very few SRH services that were financially accessible to adolescents, highlighting the need to make SRH services more accessible and available to adolescents. |
| | | | 2 | 2 | 2 | 2 | 2 | 1 | 2 | 1 | 2 | 2 | **18** | |
| 9. | Access to information and use of adolescent sexual reproductive health services: Qualitative exploration of barriers and facilitators in Kisumu and Kakamega, Kenya | Mutea, L., Ontiri, S., Kadiri, F., Michielesen, K. and Gichangi, P. (2020) | Yes | Yes | Yes | Yes | Yes | Can't tell | Yes | Yes | Yes | Valuable resource | | It provided detailed insights into the barriers adolescents face when seeking SRH services, which could assist in planning and policy development. |
| | | | 2 | 2 | 2 | 2 | 2 | 1 | 2 | 2 | 2 | 2 | **19** | |

| S/N | Title | Author (s)/Date of publication | Responses and scoring of the 10 questions on the CASP checklist | | | | | | | | | | Judgement/ Total | Reason |
|---|---|---|---|---|---|---|---|---|---|---|---|---|---|---|
| | | | Q1 | Q2 | Q3 | Q4 | Q5 | Q6 | Q7 | Q8 | Q9 | Q10 | | |
| 10 | Does Making Clinic-based Reproductive Health Services More Youth-friendly Increase Service Use by Adolescents? Evidence From Lusaka, Zambia | Mmari and Magnani (2003) | Yes | Yes | Yes | Yes | Yes | Can't tell | Can't tell | Yes | Yes | Valuable resource | | It is unclear how the authors situated themselves in the study. Also, it is unclear how issues of ethics, consent, confidentiality, and anonymity were addressed. However, the study highlighted the need for youth friendly sexual and reproductive health services for youths. |
| | | | 2 | 2 | 2 | 2 | 2 | 1 | 1 | 2 | 2 | 2 | **18** | |
| 11 | Adolescents living with HIV in the Copperbelt Province of Zambia: Their reproductive health needs and experiences. | McCarraher et al. (2018) | Yes | Yes | Yes | Can't tell | Yes | Can't tell | Yes | Yes | Yes | Valuable resource | | Although information regarding participant recruitment and author's position is unclear, the study highlighted contraceptive information and other sexual and reproductive service needs of adolescents living with HIV. |
| | | | 2 | 2 | 2 | 1 | 2 | 1 | 2 | 2 | 2 | 2 | **18** | |
| **12.** | Rights-based services for adolescents living with HIV: adolescent self-efficacy and implications for health systems in Zambia | Mburu et al. (2013) | Yes | Yes | Yes | Yes | Yes | Can't tell | Yes | Yes | Yes | Valuable resource | | Information regarding the relationship between the authors and study participants appears unclear. However, gaps in SRH service delivery of adolescents living with HIV in Zambia were identified, which could enhance a re-design of the services for quality provision of SRHS.. |
| | | | 2 | 2 | 2 | 2 | 2 | 1 | 2 | 2 | 2 | **2** | **19** | |

| S/N | Title | Author (s)/Date of publica-tion | Q1 | Q2 | Q3 | Q4 | Q5 | Q6 | Q7 | Q8 | Q9 | Q10 | Judgement/ Total | Reason |
|---|---|---|---|---|---|---|---|---|---|---|---|---|---|---|
| | | | Responses and scoring of the 10 questions on the CASP checklist | | | | | | | | | | | |
| 1<br>13 | Accessing Sexual and Reproductive Health Information and Services: A Mixed Methods Study of Young Women's Needs and Experiences in Soweto, South Africa. | Lince-Deroche et al. (2015) | Yes | Yes | Yes | Yes | Yes | Can't tell | Yes | Yes | Yes | Valuable resource | | Documentation on author's position seems unclear, however, the study need to explore support systems for adolescents. |
| | | | 2 | 2 | 2 | 2 | 2 | 1 | 2 | 2 | 2 | 2 | 19 | |
| 14. | Adolescents' Reproductive Health Problems, Service Preferences, and Accessibility. | Kimo et al. (2017) | Yes | Yes | Yes | Can't tell | Yes | Can't tell | Yes | Can't tell | Yes | Valuable resource | | The authors were unclear on their position in the study, recruitment process and documentation on data analysis appears insufficient. However, study findings revealed service delivery needs such as equipment, low-cost services and man-power needs which could assist decision makers plan acceptable adolescent SRH services. |
| | | | 2 | 2 | 2 | 1 | 2 | 0 | 2 | 1 | 2 | 2 | 16 | |
| 15. | Living as an adolescent with HIV in Zambia – lived experiences, sexual health, and reproductive needs. | Hodgson et al. (2012) | Yes | Yes | Yes | Yes | Yes | Can't tell | Yes | Yes | Yes | Valuable resource | | The study identified gaps in integrating SRH with HIV and psychosocial services, despite no clear documentation regarding the researcher's roles and positions in the study. |
| | | | 2 | 2 | 2 | 2 | 2 | 1 | 2 | 2 | 2 | 2 | 19 | |

| S/N | Title | Author (s)/Date of publication | Q1 | Q2 | Q3 | Q4 | Q5 | Q6 | Q7 | Q8 | Q9 | Q10 | Total | Reason |
|---|---|---|---|---|---|---|---|---|---|---|---|---|---|---|
| | | | Responses and scoring of the 10 questions on the CASP checklist | | | | | | | | | | | |
| 16 | Young people's perception of sexual and reproductive health services in Kenya | Godia et al. (2014) | Yes | Yes | Yes | Yes | Yes | Yes | Yes | Yes | Yes | Valuable resource | | The study was sufficiently rigorous. It revealed the opinions of young people regarding SRHS, indicating evidence linking the presence of recreational services at youth centres and general health facilities to increase access and utilisation of Adolescent SRH services. |
| | | | 2 | 2 | 2 | 2 | 2 | 2 | 2 | 2 | 2 | 2 | **20** | |
| 17 | Understanding sexual and reproductive health needs of adolescents: evidence from a formative evaluation in Wakiso district, Uganda | Atuyambe et al. (2015) | Yes | Yes | Yes | Yes | Yes | Can't tell | Yes | Yes | Yes | Valuable resource | | While it is unclear how the researchers situated themselves in the study, the study identified the adolescent problems and the need to for adolescent friendly sexual and reproductive health services. |
| | | | 2 | 2 | 2 | 2 | 2 | 1 | 2 | 2 | 2 | 2 | **19** | |
| 18. | Adolescent human immunodeficiency virus self-management: Needs of adolescents in the Eastern Cape | Adams and Crowley (2021) | Yes | Yes | Yes | Yes | Yes | Yes | Yes | Yes | Yes | Valuable resource | | The study was sufficiently rigorous. It identified the self-management needs of ALHIV, and this could help decision makers plan and develop programmes to this group of adolescents. |
| | | | 2 | 2 | 2 | 2 | 2 | 2 | 2 | 2 | 2 | 2 | **20** | |
| 19 | Preferences for accessing sexual and reproductive health services among adolescents and young adults living with HIV/ AIDs in Western Kenya: A qualitative study | Adhiambo, H. F, Ngayo, M. and Kwena, Z. (2022) | Yes | Yes | Yes | Yes | Yes | Can't tell | Yes | Yes | Yes | Valuable resource | | Although there is paucity of documentation on the relationship between the researchers and participants in terms of potential bias, the study is highly valuable because it highlights youth's preferences for accessing SRH services which could be helpful in designing responsive SRHS in the setting. |
| | | | 2 | 2 | 2 | 2 | 2 | 1 | 2 | 2 | 2 | 2 | 19 | |
| 20 | Pregnancy and STI/HIV prevention intervention preferences of South African adolescent girls: findings from | Twitty et al. (2023) | Yes | Yes | Yes | Yes | Yes | Can't tell | Yes | Yes | Yes | Valuable resource | | The authors were not clear on how they considered the relationship between the researcher and participants. However, the study is valuable in terms of exploring the STI/HIV, and pregnancy prevention preferences of young females which could assist in |
| | | | 2 | 2 | 2 | 2 | 2 | 1 | 2 | 2 | 2 | 2 | **19** | |

| | | | | | | | | | | | | | | |
|---|---|---|---|---|---|---|---|---|---|---|---|---|---|---|
| | a cultural consensus modelling qualitative study | | | | | | | | | | | | | the design of specific interventions and encourage access of SRHS by young females. |

**Scoring:**

Responses: **Q1- Q9 = yes (2); can't tell (1); No (0)**
**Q10 = Valuable resource (2); less valuable (1)**

**Judgement**

**1 - 7 = Low quality**

**8-14 = Medium Quality**

**15- 20= High quality**
